# Evidence for Action: Coverage and Access gaps in Integrated Management of Acute Malnutrition in Kapilvastu district of Nepal

**DOI:** 10.1101/2025.08.01.25332576

**Authors:** Bibek Kumar Lal, Sujay Nepali, Roshna Maharjan, Ashish Timalsina, Melkamnesh Alemu Nigussie, Phulgendra Prasad Singh, Manisha Katwal

**Author notes:** Corresponding author: /.

## Abstract

Wasting remains a critical public health concern in Nepal, with many children unable to access timely treatment inspite of national programmatic efforts. Despite implementation of the program since 2012/13 to address acute malnutrition, the evidence about the program coverage and factors supporting as well as hindering the program coverage is lacking in Kapilvastu district. The SQUEAC approach was implemented in three stages using a mixed-method design. Stage I involved qualitative data collection through key informant interviews, focus group discussions, and direct observations in 18 locations. Stage II tested a hypothesis about proximity and coverage via small-area surveys adopting active adaptive and door-to-door case finding. Quantitative data from DHIS-2 and IMAM registers were analyzed for trends and performance indicators, while qualitative data were thematically analyzed and triangulated across participants, types and methods. A Barriers, Boosters, and Questions (BBQ) scoring exercise informed prior coverage estimation. Upon validation and confirmation of the hypothesis, wide area survey was conducted to estimate the coverage of the program. The assessment identified 14 boosters, ranging from active involvement of trained Female Community Health Volunteers (FCHVs) and Health Workers (HWs), strong community trust in frontline workers, to the consistent availability of Ready to Use Therapeutic Food (RUTF) in health facilities. Similarly, 24 barriers were identified which ranged from poor economic conditions, limited awareness of malnutrition among families, to cultural beliefs that prioritize traditional healers over modern healthcare. The other barriers included logistical challenges such as distance to OTCCs, stockouts of essential commodities, and high turnover rates among trained health workers. The assessment validated the hypothesis “Coverage of the program is high in communities having low concentration of DAG and low in communities having high concentration of DAG in both rural and urban context”. Upon validation of the hypothesis, wide area survey was conducted which estimated the coverage at 22.9%, with a confidence interval of 14.3% to 34.8%. The point coverage of the program was found to be 6.67% while the period coverage, which accounts for both enrolled and recently recovered cases, was found to be 12.5%. The IMAM program in Kapilvastu faced challenges on both service delivery side as well as service seeking sides. On the service delivery side, it faced systemic challenges. These included inadequate capacity building or refresher to human resources, unavailability of IMAM guidelines and protocols, stock out of RUTF, and deviation from IMAM protocols. It was found that the children enrolled in IMAM program who received RUTF for treatment, were distributed super cereal once they fell in MAM category, deviating from IMAM protocol which strictly instructs to provide RUTF until the case is completely recovered.

## INTRODUCTION

Wasting is a critical public health concern significantly contributing to increased child morbidity and mortality. Children with severe wasting are 11 times more likely to die from common childhood illnesses compared to the well-nourished children. (1) Although the prevalence of wasting declined from 10% in 2016 to 8% in 2022 (2), Nepal falls significantly short of achieving its national targets of reducing acute malnutrition below 5% as the goal set in the Second-Long Term Health Plan (1997-2017) and the Multi Sector Nutrition Plan (MSNP I and II). Sustainable Development Goals (SDG) have set even more ambitious target of less than 5% by 2025, and to 4% by 2030. (3)

To address acute malnutrition, Government of Nepal implemented IMAM program, previously known as Community Management of Acute Malnutrition (CMAM) program in 2008/9, expanding it from 5 districts to 38 districts by 2020. (4) Furthermore, the Government of Nepal endorsed the Comprehensive Nutrition Specific Intervention (CNSI) package and implemented across all 77 districts of Nepal. Despite these efforts, many acutely malnourished children remain undetected within the community hindering them to access timely and appropriate treatment.

The IMAM program addresses wasting through a community-based approach, including nutrition education, therapeutic feeding at Outpatient Therapeutic Care Centers (OTCCs) for Severely Acutely Malnourished (SAM) cases without medical complications, those children with medical complications are managed at Inpatient Therapeutic Care Centers (ITCCs). Moderate wasting is addressed through counselling mothers/caretakers on Infant and Young Child Feeding (IYCF) practices, and hygiene promotion.

The program has been implemented in Kapilvastu since 2012/13 to address the acute malnutrition and improve the nutrition outcome of the district. However, the evidence on program coverage, and factors supporting and hindering the coverage are still lacking. Therefore, this coverage assessment plays a crucial role in generating the evidences on the factors boosting up or pulling down the coverage. At the same time, it also provides the coverage estimate, point coverage and period coverage of the program.

## METHODS

This assessment strictly followed the standard coverage assessment methodology called Semi Quantitative Evaluation of Access and Coverage developed by Valid International, FHI 360 / FANTA, UNICEF, Concern Worldwide, World Vision International, Action Against Hunger, Tufts University, and Brixton Health. The necessary tools were also adopted from the tools available in coverage monitoring network which were adapted in the context of IMAM program implementation in Nepal. The methodology is three-stage procedure:

1. Stage I: Identification of high and low coverage areas
2. Stage II: Validation of hypothesis
3. Stage III: Wide Area Survey/ Coverage Estimate

### Stage I: Identification of High and Low Coverage areas

Stage I identifies the areas of high and low coverage through quantitative data analysis of routine program data. This analysis was done basically to inform the assessment team regarding the admission pattern, MUAC on admission and exit, discharge outcomes, length of stay, relation between case identification and seasonality. Qualitative data was collected via KII, FGDs, and direct observations. The qualitative data is collected to identify the boosters supporting the effectiveness of the program and barriers hindering the program coverage and access.

#### Tools

Respondent-oriented key informant interview guides, FGD guides and direct observation checklists were developed. These tools have been previously tested during the same assessment conducted in other districts and hence the tools were adapted modifying the questions according to the current context of the program implementation.

#### Respondents

The respondents for this assessment were mother’s or caretakers of various categories of cases as defined by IMAM guideline and the stakeholders involved in the program. Basically, in this assessment the following were the respondents

- Mother or caretaker of SAM children who are enrolled in the program
- Mother or caretaker of SAM children not covered in IMAM program
- Mother or caretaker of SAM children who defaulted from IMAM program
- Nutrition focal person at OTCC
- Female Community Health Volunteer
- OTCC In-charge
- Representative of a community-based organisation
- Community leader (ward representative)
- Teacher
- Mother of cured child
- Mothers of under five children
- Nutrition focal person at health office

Unlike in other conventional studies, the sample size or the respondents for the collection data is not predetermined for the assessment. The sample size is determined as the stages progress.

Stage I strictly adheres to the principle of saturation and triangulation of data by source method and location. Hence, the qualitative data in Stage I is collected until it achieves the saturation point. For further validation of data, FGDs were conducted with FCHVs, and mothers of under 5 children using FGD guides and direct observation at OTCCs. The qualitative data were collected from 18 different locations of Kapilvastu district.

### Data Management and Analysis

Qualitative data collected was synthesized in the form of Barriers, Boosters and Recommendations (BBR) exercise. All the information on barriers, boosters, and recommendations were synthesized in 8 thematic areas out of which 6 belonged to six WHO building blocks (5), and the rest 2 areas were community engagement, and gender and social inclusion (GESI). Each of the barriers and boosters was thoroughly synthesized, and documented by source, method and location^6^. The sources, methods and locations were denoted with symbols so that there is evidence of various sources, methods, and locations for increasing the weight and validity of each barrier and booster associated with hindering and enhancing the effectiveness of program as well as the program coverage and access. Later, the synthesized Barriers and Boosters were weighed using weighted score and simple score. In weightage scoring each of the barriers and booster was given scoring between 1 to 4 whereas in simple scoring, each barrier and booster was given equal score i.e.4. (6)

### Stage II: Setting a hypothesis

This stage is for setting the hypothesis and testing the same. Based on the level of impact on the effectiveness, coverage and access of program, the hypothesis was set which stated, **“Coverage of the program is high in communities having low concentration of DAG and low in communities having high concentration of DAG in both rural and urban context”.** During this stage, cases were identified following active-adaptive case finding in rural context, and door to door in urban context, deploying small area survey. The case finding was done in 12 different locations in both the context, urban and rural. Three locations each with high concentration of disadvantage groups (Muslims / Dalits), and 3 locations each with low concentration of disadvantage group were selected.

#### Tools

Separate standard questionnaire for covered and uncovered cases were adopted from coverage monitoring network. The questionnaires were administered to the mothers or caretakers of covered cases, i.e. SAM cases enrolled and currently under treatment, and the cases enrolled in the program who have graduated from SAM to MAM. The uncovered questionnaire was administered to the mothers or caretakers of children who are SAM but not enrolled in the program at the time of assessment. The entire case finding process included the children between 6 to 59 months of age.

### Stage III: Wide Area Survey

This stage is also called a wide area survey or likelihood survey. Before moving to this stage, the prior was developed based on the scoring from the Barriers, Boosters, and Questions (BBQ) exercise. Prior mode provides a guesstimate about the program coverage which informs the assessment team that the coverage would be somewhere around the prior mode. This is calculated by taking the average of simple and weighted.

The sample for wide area survey or likelihood survey was calculated using the alpha prior and beta prior and the uncertainty level. Since this assessment was conducted for the first time in the district, the uncertainty level of 25% is taken. Using the standard table for 25% given the SQUEAC guideline, Alpha prior of 11.1, Beta prior of 20.6, and precision of 12 was plugged-in in Bayesian scale for suggested sample (n) which was 30. Here the calculated sample size was the number of SAM cases (n) required for wide area survey. This required to be translated into the minimum number of villages that needed to be sampled to achieve the required sample size. The required number of villages was calculated using the following formula: **n _villages-=_n/ [average village population _all ages_ * percentage of population _6-59months_/100 *SAM prevalence/100] *100.** (6)

The case finding is done via active adaptative approach in rural and door-to-door approach in urban context. Case findings were done through anthropometric measurement using mid-upper arm circumference (MUAC) tape and checking the bilateral pitting edema.

#### Tools

The standard questionnaire, adopted and adapted from coverage monitoring network, was used to collect qualitative information on reasons for enrolling and not enrolling in the program.

Two different types of questionnaires were used, one for mothers/caretakers of covered or enrolled SAM cases and another for SAM cases out of program meaning that the cases were identified as SAM during assessment but were out of treatment (not covered by the program). Tally sheet of coverage monitoring network was also used to track the date of survey and number of cases identified.

### Ethical Statement

Ethical approval for this study was obtained from the Nepal Health Research Council (NHRC) under the approval number 543_2024. Participation in the study was voluntary, and informed written consent was obtained from all participants prior to interviews and focus group discussions. All data were anonymized to ensure participant privacy.

## RESULTS

The results are presented by Stage as this assessment is a 3 phase procedure where the result of one procedure leads to another stage.

**Stage 1:** The quantitative data was analyzed for admission over time, MUAC at admission, discharge over time and length of stay and based on the quantitative analysis, areas of low and high coverage were identified to identify the reasons for high and low coverage of the program.

### Quantitative Findings

#### Admission overtime

The data for admission over time showed the inconsistencies in the number of children admitted in the OTCCs. The admissions were consistent in the months mid July to mid to mid October to some extent, which declined significantly in mid October-mid November and mid January-mid February. Mid July to mid September months fall in the monsoon season, a period associated with high prevalence of diseases such as diarrhoea and malaria, which often lead to malnutrition. The decline in admissions during mid October-mid November may be attributed to the festival season (Dashain and Tihar), when food availability generally improves for many households.

Conversely, mid March to mid May months, which fall during the dry season, often see a rise in health issues related to seasonal changes, such as respiratory infections and water scarcity-related illnesses, impacting the overall nutritional status of children. Hence the identification and admission in OTCC increased.

**Figure 1:**
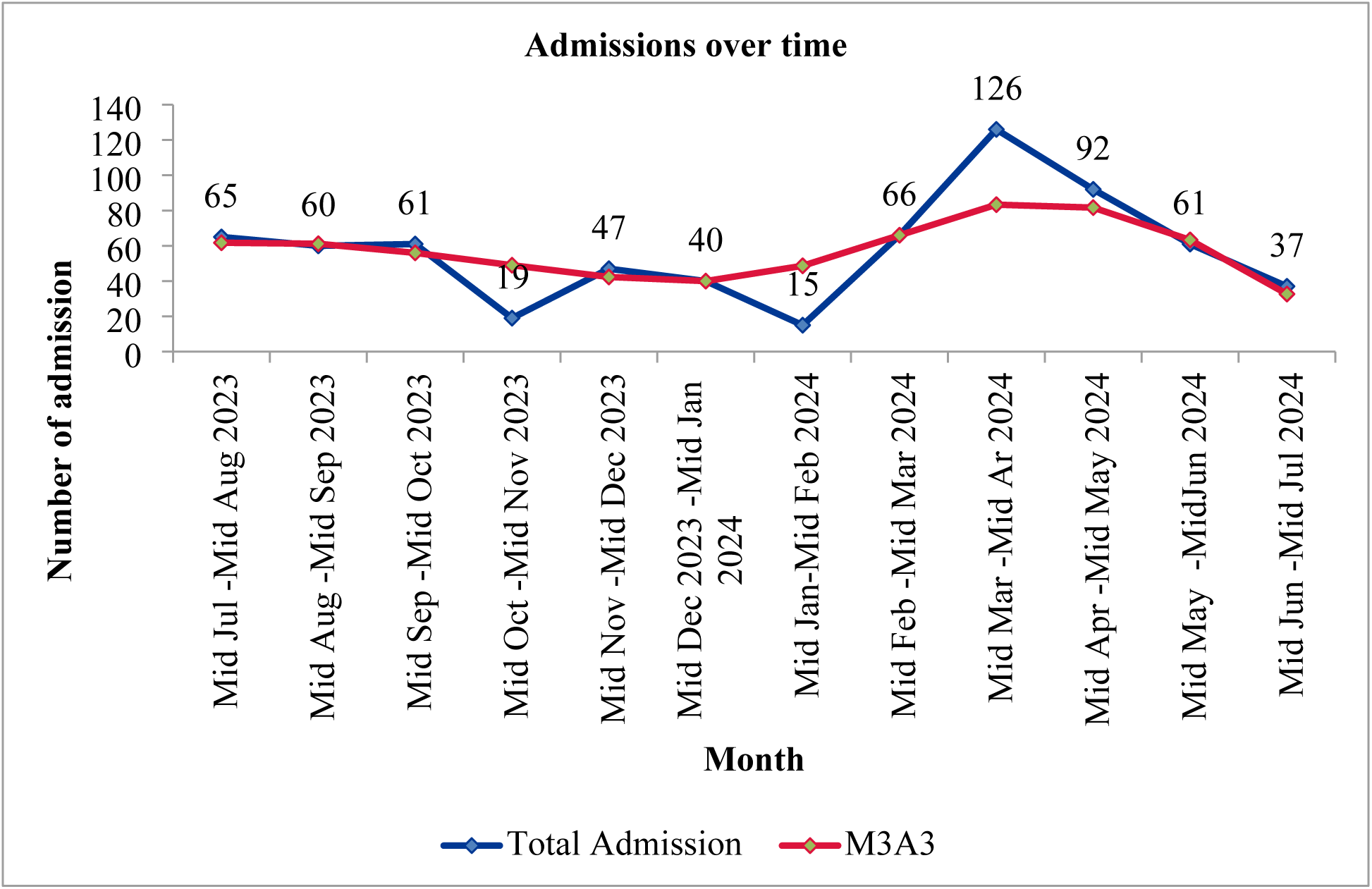
Admissions over time

### MUAC at admission

The data on MUAC at admission shows that MUAC peaks at 115 mm and 110 mm, which tells that most of children are admitted to OTCCs either when their MUAC is 115 mm or when it is 110 mm. Since the cases were also enrolled in the program using z-score, MUAC values of more than or equal to 115 mm were also observed. However, there might have been wrong admissions by MUAC, which cannot be clarified from the graph. The same figure also shows that children with MUAC measurements of less than 115 mm as well as less than 125 mm were mentioned, without pointing the exact MUAC measurement in the registers. Additionally, cases with MUAC measurements as low as 80 mm, 95 mm, and 100 mm were also recorded. This highlights late detection and delayed admission of cases into the program. Such delays place children at a significantly higher risk of health complications and vulnerability.

**Figure 2:**
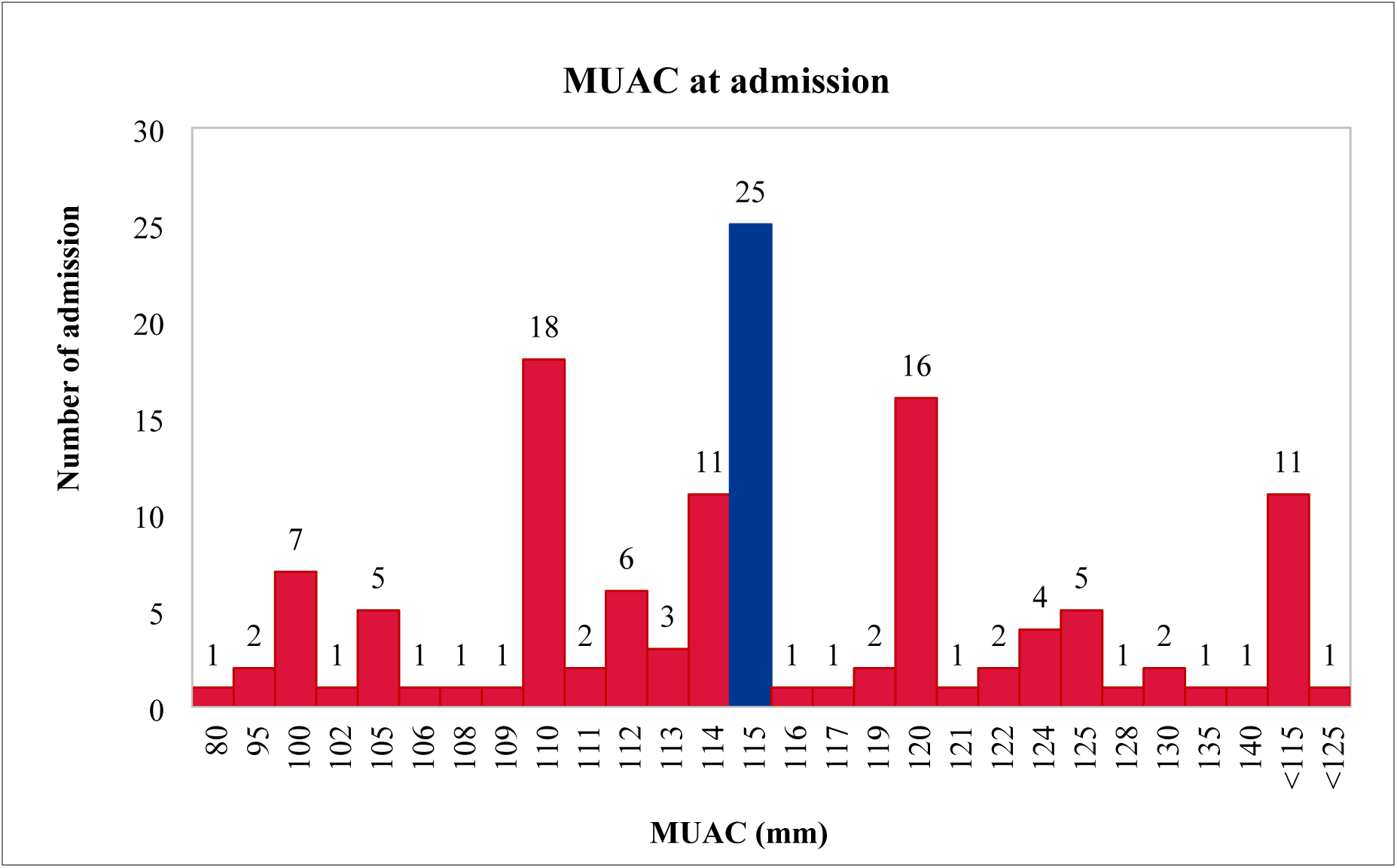
MUAC at admission

### Discharge Over time

The data for discharge over time reveals that recovery rate for only six months meet the Sphere standard which is >75%. Similarly, the defaulter rate exceeds the Sphere standard of <15%

**Figure 3:**
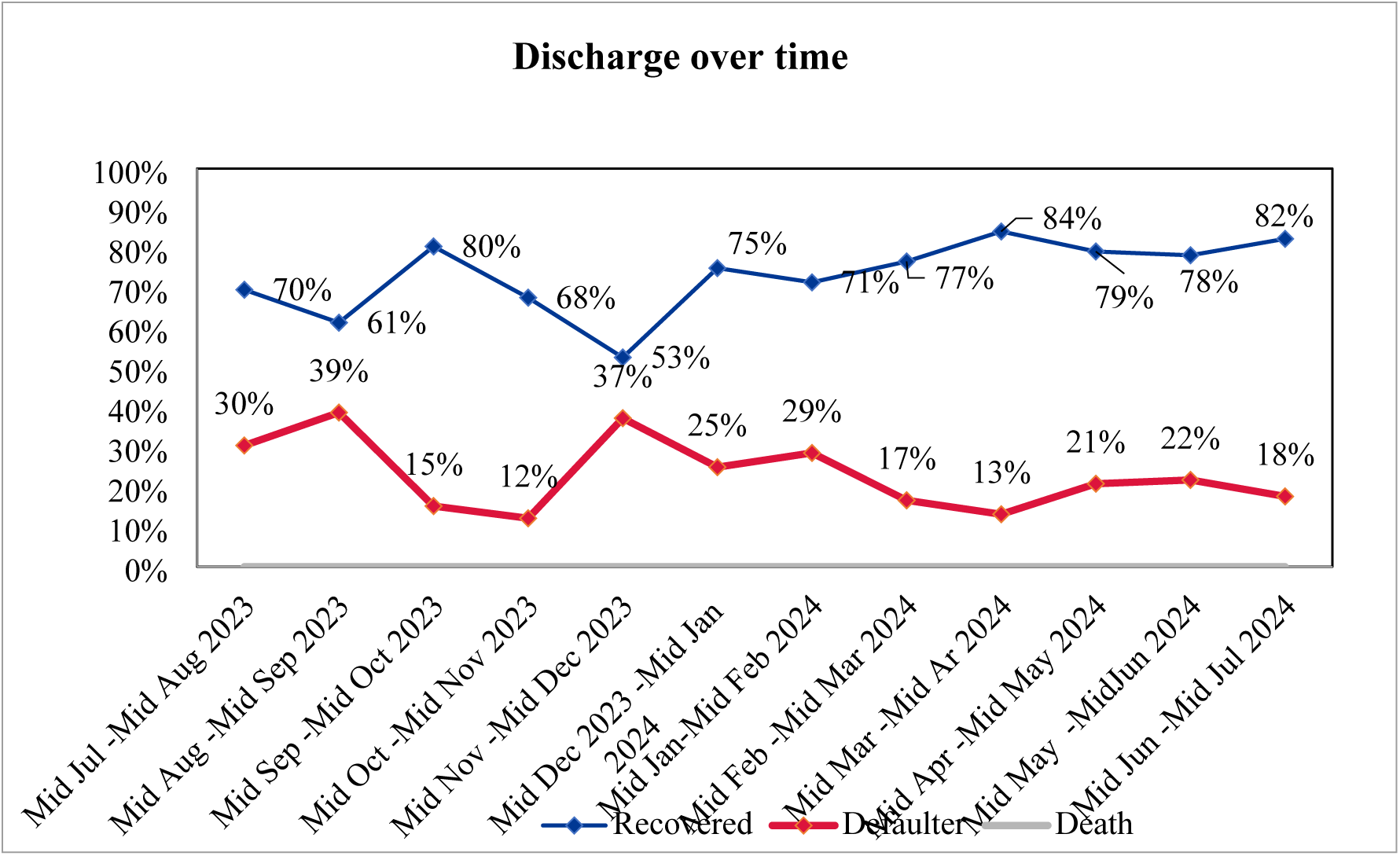
Discharges over time

### Length of Stay (week) before discharge as recovered

The median length of stay in the program for children ‘discharged as cured’ was 9 weeks. This means that 50% of children were discharged within 9 weeks or less. The minimum length of stay for recovery was 3 weeks, suggesting that some children responded quickly to treatment.

However, the maximum length of stay extends to 32 weeks. Such prolonged stays may indicate challenges, such as cases with severe co-morbidities, late detection of malnutrition, or issues with treatment adherence and follow-up.

The graph shows that the median number of visits before defaulting from the program was 4 visits, meaning 50% of the children default after 4 or fewer visits. However, there were cases where children defaulted after just one visit, indicating that some come for admission but fail to return for any follow-up visits.

**Figure 4:**
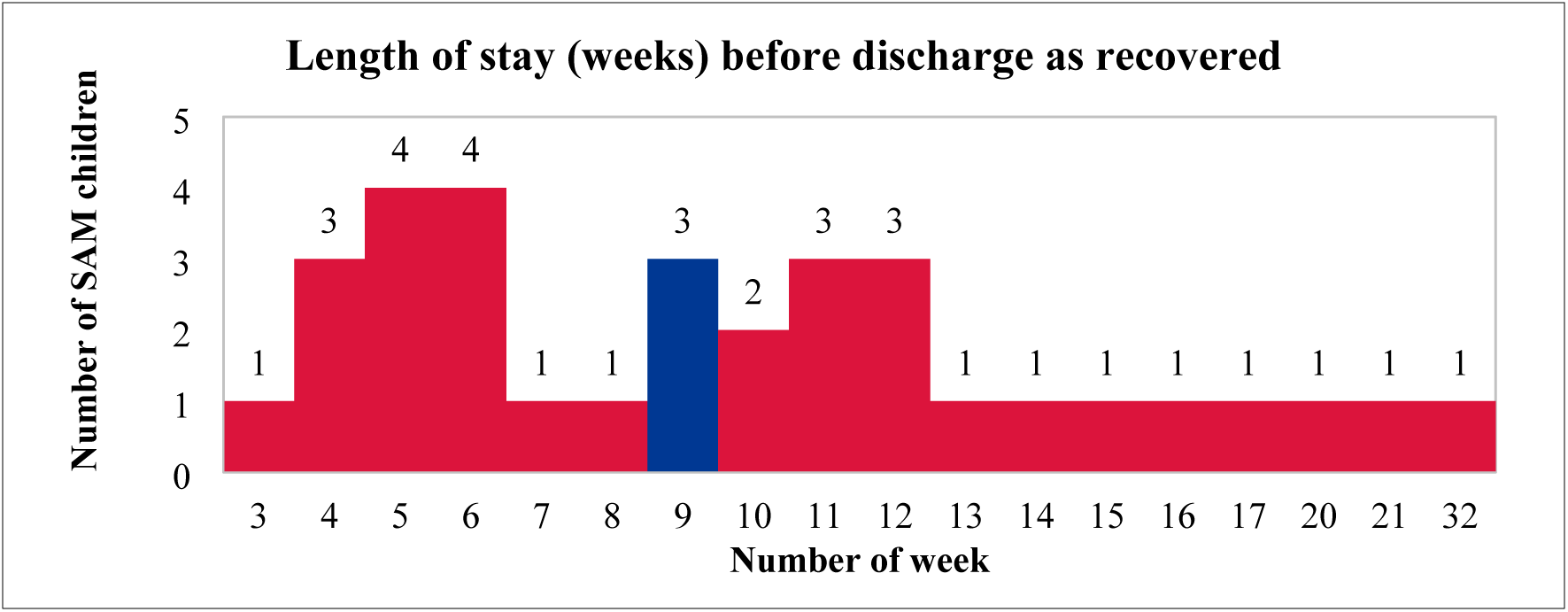
Length of stay (week) before discharge as recovered

**Figure 5:**
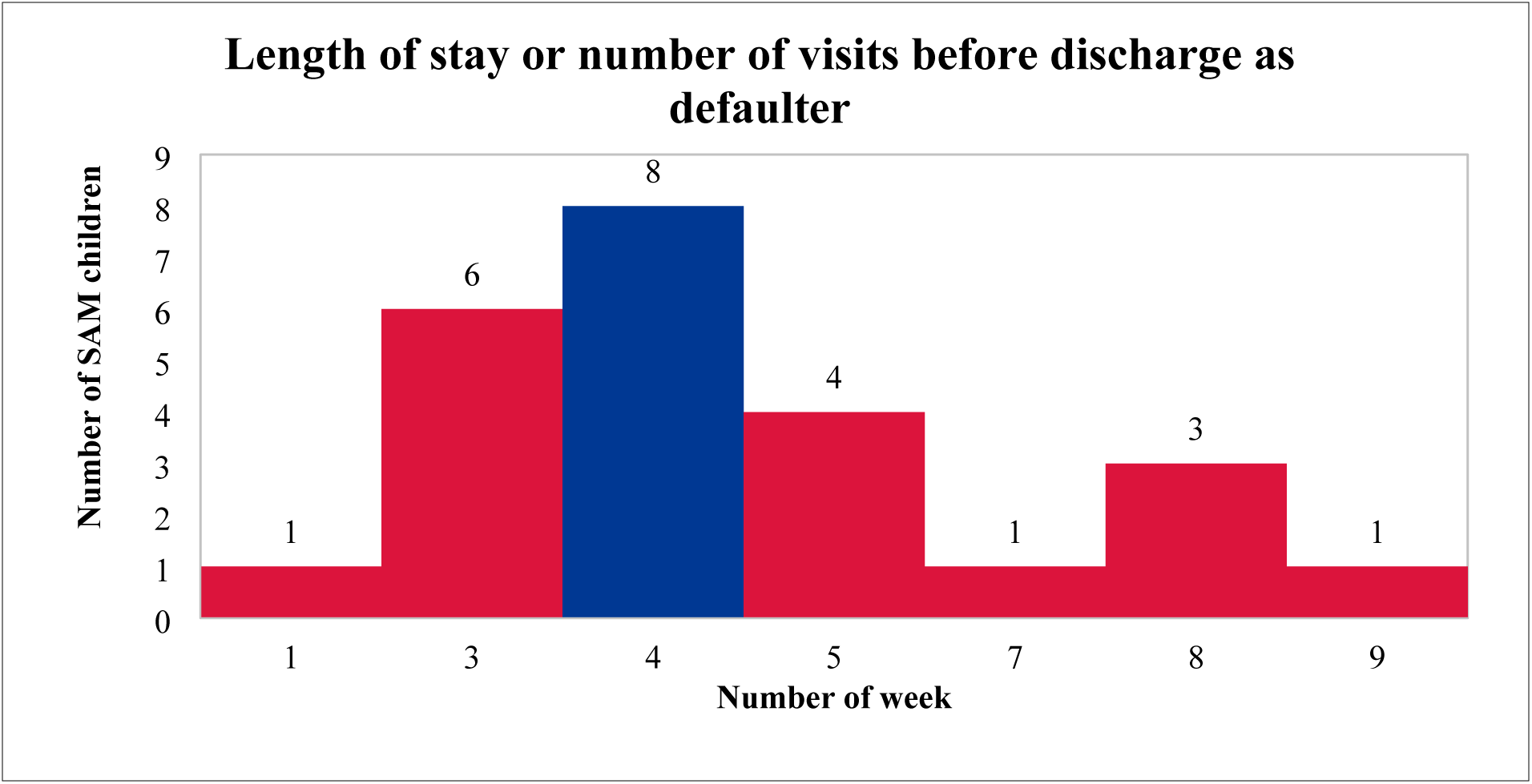
Length of stay or number of visits before discharge as defaulter

### Qualitative finding

Based on the quantitative data analysis, 18 different locations were selected for identification of factors associated with low and high coverage. Fourteen boosters and twenty-four barriers were identified. Barriers were identified for both service delivery side as well as service uptake side, which primarily included service delivery and uptake. Cultural restrictions, particularly in Muslim communities, require women to obtain permission from male family members before accessing healthcare services. Language barriers at service delivery sites further hinder access. In addition, unwelcoming behavior from some health workers discourages service seekers, and frequent stockouts of RUTF limit treatment availability. The long distances between OTCCs and beneficiaries’ homes, combined with a general lack of awareness among mothers about the program, also contribute to low service utilization.

The boosters mainly included the active involvement of trained Female Community Health Volunteers (FCHVs) and Health Workers (HWs), strong community trust in these frontline workers, and the consistent availability of RUTF in health facilities. Local initiatives such as nutrition allowances for malnourished children and awareness campaigns, along with large number of OTCCs, are also contributing to the district’s efforts to identify the cases and bring them into the program catchment and provide timely treatment.

The BBQ exercise evaluated 14 boosters and 24 barriers using both a simple scoring and a weighted scoring method, as presented in the following table. In the simple scoring method, each booster and barrier was assigned an equal score, if all factors influenced the program’s effectiveness equally. In contrast, the weighted scoring method assigned scores based on the perceived level of impact that each factor had on the program’s outcomes.

**Table 1:** Barriers and Boosters Weighing.

| S.N. | Positive factor<br>(Booster) | Weighing |  | Negative factor<br>(Barrier) | Weighing |  |
| --- | --- | --- | --- | --- | --- | --- |
|  |  | Simple | Weighted |  | Simple | Weighted |
| 1 | Active FCHVs<br>(screening,<br>counselling) | 4 | 2.5 | Poor economic<br>condition of<br>households | 4 | 3 |
| 2 | Trained FCHVs and<br>HWs in CNSI | 4 | 3.5 | No / less awareness<br>of community on<br>malnutrition | 4 | 3.5 |
| 3 | CBOs/NGOs/partner<br>working in nutrition<br>available | 4 | 3 | Superstition (faith on<br>traditional healer) | 4 | 2 |
| 4 | Increasing trust of<br>mothers /community<br>in FCHVs and HWs | 4 | 2 | Cultural barriers | 4 | 2.5 |
| 5 | Responsible Health<br>Workers | 4 | 2 | No/limited decision<br>power of mothers<br>(dominant in-<br>laws/male) | 4 | 3.5 |
| 6 | Adequate stock level<br>of RUTF maintained<br>in HF's | 4 | 3.5 | Negligence in<br>treatment from<br>mothers/family | 4 | 2.5 |
| 7 | Supportive family | 4 | 2.5 | Long distance from<br>OTCCs/HF's | 4 | 2.5 |
|  |  | Simple | Weighted |  | Simple | Weighted |
| 8 | Adequate number of OTCCs established in the district (44) | 4 | 2 | No designated room for OTCC | 4 | 3 |
| 9 | Supportive linkage between OTCCs (supported to circulate RUTF) | 4 | 2.5 | RUTF shortage / stockout | 4 | 2 |
| 10 | Presence of influential people (*Maulana*, political leaders) | 4 | 1.5 | Unavailability of updated/IMAM guidelines | 4 | 3 |
| 11 | Nutrition allowance for SAM children (*Banganga*, *Shivraj* R/Ms) | 4 | 1.5 | Commodities shortage / stockout | 4 | 3 |
| 12 | Malnutrition free ward declaration campaign (Ward 5 & 8 of *Banganga* municipality) | 4 | 2 | High turn-over of trained HWs | 4 | 1.5 |
|  |  | Simple | Weighted |  | Simple | Weighted |
| 13 | MAM cases covered by other project (Super Cereal) | 4 | 2.5 | Unwillingness of HWs to admit SAM children (from outside of their catchment area) | 4 | 1 |
| 14 | Provision of incentive for FCHVs | 4 | 2 | Behavioural concerns of Health Workers | 4 | 2.5 |
| 15 |  |  |  | Irregularity in service delivery | 4 | 1 |
| 16 |  |  |  | Language barrier | 4 | 2 |
| 17 |  |  |  | Refusal of RUTF by SAM child | 4 | 2 |
| 18 |  |  |  | Improper storage of RUTF (tin trunk used for other commodities/sharing, damaged RUTF) | 4 | 1.5 |
| 19 |  |  |  | RUTF sharing with other children and adults | 4 | 2.5 |
|  |  | Simple | Weighted |  | Simple | Weighted |
| 20 |  |  |  | No budget for MGM<br>and mobilisation of<br>FCHVs | 4 | 3 |
| 21 |  |  |  | Inactive FCHVs<br>(SAM child near<br>FCHV's house) | 4 | 2 |
| 22 |  |  |  | Poor health seeking<br>behaviours | 4 | 2.5 |
| 23 |  |  |  | Absence of FCHV<br>(FCHV abroad, no<br>new recruitment due<br>to political pressure) | 4 | 2 |
| 24 |  |  |  | Limited<br>transportation to go<br>to HFs/OTCCs | 4 | 2 |
|  |  | 56 | 33 |  | 96 | 56 |
|  |  | 56 | 33 |  | 4 | 44 |

#### Stage 2

A hypothesis was set based on the barriers and boosters identified during stage 1 and scoring done during BBQ exercise. This hypothesis was confirmed through a small area survey.

Hypothesis was set around awareness which stated, **“*Coverage of the program is high in communities having low concentration of DAG and low in communities having high concentration of DAG in both rural and urban context”*.**

An active adaptive case-finding approach was implemented to test the hypothesis. A total of three SAM cases were identified in both the areas/locations with high and low concentration of DAG. In rural context, a total of 3 cases among which 2 covered cases (bilateral pitting oedema and/or MUAC <115mm and currently in OTCC for treatment) were identified in the communities with low concentration of DAG. Similarly, 4 cases were identified among which 1 covered case was identified in areas/locations with high concentration of DAG. Similarly, in the urban context, a total of 4 SAM cases were identified in areas/locations with low concentration of DAG, and 5 cases were identified in areas/locations with high concentration of DAG. Among 4 cases in the communities with low concentration of DAG, 3 were enrolled in the program and were receiving treatment at the OTCCs. Similarly, a total of 5 uncovered SAM cases (bilateral pitting oedema and/or MUAC <115mm and not currently in OTCC for treatment) were identified in areas/locations with high concentration of DAG. The decision rule was satisfied in both rural and urban context and thus the hypothesis was confirmed and validated.

#### Stage 3

Since the hypothesis was confirmed and validated in stage II, Stage III proceeded for likelihood or wide area survey. Stage 3 identified a total of 32 SAM cases during the survey, out of which only 2 cases (6%) were enrolled in the treatment program, while another 2 cases (6%) were in the recovery phase. Among the recovering cases (MUAC >115mm but still under-going OTCC for treatment (taking RUTF); has not yet meet the IMAM program’s discharge criteria), one was male, and another was female child. Alarmingly, 28 out of 32 identified cases (88%) were not enrolled in the program, leaving them untreated and at significant health risk. Notably, 24 (86%) of these uncovered cases were female, while only 4 (14%) were male, indicating a potential gender disparity in access to care. Additionally, 8 of the uncovered cases had previously been enrolled but had relapsed at the time of the survey.

Caretakers of defaulting children cited several reasons for discontinuing treatment. Some children disliked the taste of RUTF or experienced side effects such as vomiting and nausea. Other caretakers faced personal challenges, such as pregnancy, which limited their ability to take the child to the health facility, while some lacked sufficient family support for childcare.

Additionally, distrust in health facilities emerged as a barrier, with some mothers perceiving the center as only offering basic health services.

Similarly, among the 22 caretakers who had never sought SAM treatment for their children at OTCCs, several barriers were reported. These included a lack of awareness about OTCC services, a preference for private clinics over public health facilities, and dissatisfaction with treatment at OTCCs - some mothers expected RUTF but were instead provided with “Super cereal.” Many also cited a lack of family support as a significant challenge.

These findings highlight critical gaps in SAM program coverage and retention, with low enrolment, high default rates, and gender disparities demanding urgent attention.

### Coverage Estimation

The wide-area survey identified 32 SAM cases. Among these, 2 cases (C_in_) were covered in the program, 2 cases (R_in_) were recovering within the program, and 28 cases (C_out_) were uncovered. Using a Bayesian analysis with a prior coverage estimate of 34.3%, the updated overall coverage estimate was determined to be 22.9% with a confidence interval of 14.3% to 34.8%. This interval suggests that the true coverage is likely within this range. However, point and period coverage are calculated using the below formula.

**Point Coverage** = Covered case (C_in_)/ {Covered case (C_in_) + Uncovered cases (C_out_)} = 2/(2+28)*100%=2/30*100% = 6.67%

**Period Coverage** = Covered case (C_in_) + Recovering case (R_in_) / Covered case (C_in_) + Recovering case (R_in_) + Uncovered cases (C_out_) = (2+2)/(2+2+28)*100%=4/32*100% = 12.5%

### Coverage Estimate using Bayesian Scale

**Figure 6:**
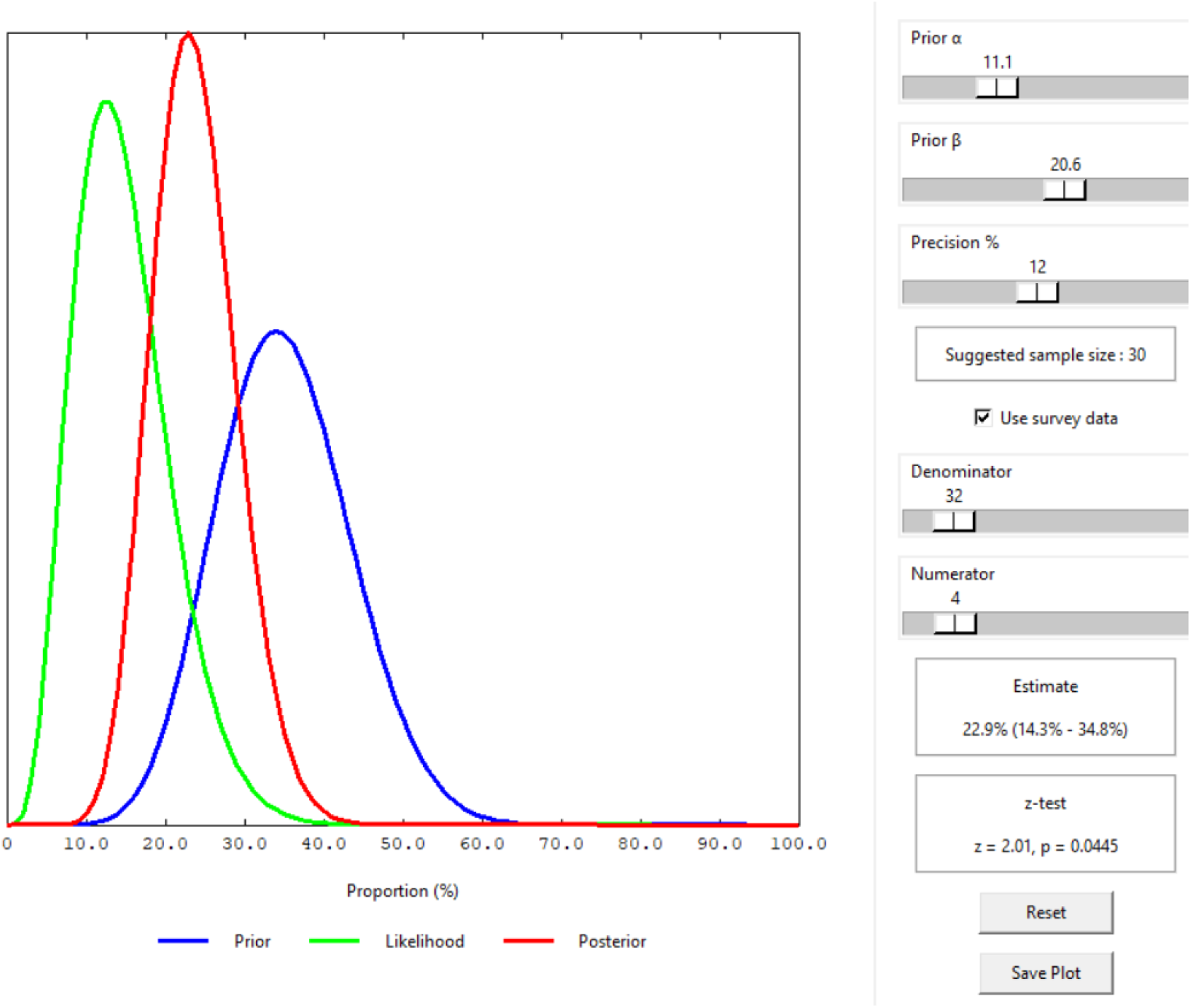
Coverage estimate using Bayesian scale

## Discussion

Kapilvastu is one of the early implementation districts for the IMAM program, which commenced in 2012. Despite its long-standing implementation, there has been no documented evidence regarding its coverage. Given the absence of systematic data, it was essential to conduct a coverage assessment to evaluate program reach and effectiveness. This assessment aimed to fill this gap by identifying areas of high and low coverage and provide an overall estimate of program uptake.

The assessment team analyzed data from OTCC registers (HMIS 2.6) and DHIS 2, revealing several gaps in record-keeping at health facilities. The poor documentation in the IMAM recording and reporting tools were also found in the various studies conducted in Kenya (7), Democratic republic of Congo (8), Somalia (9), Zambia (10). These findings highlight significant challenges in program monitoring and service delivery.

During the coverage assessment, 32 cases of SAM were identified. Of these, only 2 were enrolled in the program, 2 had recently recovered from SAM and were in the recovery phase, while the remaining 28 had not received any treatment. Based on these figures, the point coverage, which reflects the proportion of currently enrolled SAM cases, was 6.67%. The period coverage, which accounts for both enrolled and recently recovered cases, was 12.5%. Using a Bayesian statistical approach to adjust for data gaps and biases, the overall estimated coverage was calculated to be 22.9% (95% CI: 14.3% - 34.8%). This means that, when considering statistical adjustments, 22.9% of SAM cases in the district were likely covered by the program, though the actual coverage could range between 14.3% and 34.8%.

The low coverage can be attributed to multiple barriers affecting both service delivery and uptake. Cultural restrictions, particularly in Muslim communities, require women to obtain permission from male family members before accessing healthcare services. The cultural barriers hindering the health care access among Muslim community is also revealed in a narrative review conducted by John Hopkins University on Barriers to Healthcare barriers among Muslim women. (11) Language barriers at service delivery sites further hinder access^11^. Stock out of RUTF has been one of the compounding factors for smooth service delivery which was also reported in the coverage assessment done in Kenya. (12) The SQUEAC conducted in Democratic Republic of Congo, also found that stock out of RUTF along with other essential drugs in the health facilities decreased the trust among beneficiaries regarding program. (8) One of the strong factors that affected the coverage is the awareness among mothers and community which was also observed in SQUEAC conducted in Kenya (12) and Zimbawe. (10)

Capacity building of the frontline health workers is one of the strong determinants that helps to increase the coverage and access of the IMAM program.(7) Active engagement of FCHVs plays a pivotal role in service uptake and acceptance of the program by the communities, which was also revealed in similar assessment performed in Zimbawe.(10) The presence of an adequate number of OTCCs across the district contributes in increasing in access to the services which was also observed in the studies conducted in Nepal (13) and Kenya.(12) Likewise, the support from influential community leaders, and municipal funding for nutrition allowances for malnourished children, have further reinforced program sustainability. Such findings were also noted in from the studies conducted in Democratic Republic of Congo (8) and Kenya (7)

The assessment also highlighted key concerns regarding program adherence such as the practice of providing Super Cereal to recovering cases, and distribution of RUTF in simplified approach.

This raises concerns about adherence to program protocols. Issues affecting adherence to program protocols were also found in studies conducted in other countries.

## Conclusion

The SQUEAC assessment in Kapilvastu revealed both systemic and service uptake limitations. Systemic challenges were identified across all six building blocks of the WHO health system framework, as well as in community engagement and gender equality and social inclusion (GESI). To improve service uptake and increase access and coverage of the program, these systemic barriers must be addressed. In parallel, greater emphasis should be placed on awareness-raising and behavior change interventions, to enhance community participation, demand for services as well as service delivery. Additionally, strengthening the capacity of frontline health workers is a must to ensure the adherence to program protocol and quality service delivery.

## Data Availability

The data set has been attached in the supporting information with this submission.

## Acknowledgments

The authors would like to express their sincere gratitude to the Family Welfare Division for facilitating coordination with the Provincial Health Directorate, District Health Offices, and local governments to support the successful implementation of this assessment. We also extend our heartfelt thanks to UNICEF for their support throughout the process. Additionally, we are grateful to all stakeholders working in the field of nutrition, including local organizations, health offices, Health Workers, Female Community Health Volunteers, and especially the mothers who generously shared their time and insights during interviews and focus group discussions.

## References

1. Severe Wasting An Overlooked Child Survival Emergency.; 2022. Accessed April 21, 2025. https://www.unicef.org/child-alert/severe-wasting

2. Nepal Demographic and Health Survey 2022 Ministry of Health and Population New ERA Ministry of Health and Population.; 2023. www.DHSprogram.com.

3. Nepal’s Sustainable Development Goals, National Planning Commission, Kathmandu.; 2023.

4. Haag KC, Sharma A, Parajuli KR, Adhikari A. experiences_of_the_integrated_management_of_acute_malnutrition_imam_programme_in_ne pal_from_pilot_to_scale_up. Field Exch. 2020;(63). Accessed April 10, 2025. www.ennonline.net/fex

5. Monitoring the Building Blocks of Health Systems: A Handbook of Indicators and Their Measurement Strategies. World Health Organization; 2011.

6. Myatt M, Health B. Semi-Quantitative Evaluation of Access and Coverage (SQUEAC)/ Simplified Lot Quality Assurance Sampling Evaluation of Access and Coverage (SLEAC) Technical Reference THE SCIENCE OF IMPROVING LIVES.; 2012. www.fantaproject.org

7. County TB. SEMI QUANTITATIVE EVALUATION OF ACCESS AND COVERAGE (SQUEAC) SURVEY FINAL REPORT.

8. Semi-Quantitative Evaluation of Access and Coverage (SQUEAC).

9. Com/ J. SQUEAC COVERAGE SURVEY REPORT OF WORLD VISION OUTPATIENT THERAPEUTIC (OTP) AND TARGETED SUPPLEMENTARY FEEDING PROGRAMS (TSFP) IN LUGHAYA-SOMALILAND, EYL-PUNTLAND AND DOLOW-SOUTH CENTRAL, SOMALIA.

10. 1201-zmb_squeac_report_final.

11. Tackett S, Young JH, Putman S, Wiener C, Deruggiero K, Bayram JD. Barriers to healthcare among Muslim women: A narrative review of the literature. Womens Stud Int Forum. 2018;69:190–194. doi: 10.1016/j.wsif.2018.02.009

12. MARSABIT COUNTY SEMI-QUANTITATIVE EVALUATION OF ACCESS AND COVERAGE (SQUEAC) ASSESSMENT REPORT.; 2023.

13. SAPTARI DISTRICT SQUEAC INVESTIGATION 2 SAPTARI DISTRICT SQUEAC INVESTIGATION 3 ACKNOWLEDGEMENTS.; 2013.

